# Risk of hospitalization and risk of death for healthcare workers with COVID-19 in nine European Union/European Economic Area countries, January 2020–January 2021

**DOI:** 10.1101/2021.03.01.21252651

**Authors:** Lisa Ferland, Joana Gomes Dias, Carlos Carvalho, Cornelia Adlhoch, Carl Suetens, Julien Beauté, Pete Kinross, Diamantis Plachouras, Favelle Lamb, Tuula Hannila-Handelberg, Massimo Fabiani, Flavia Riccardo, Joël Mossong, Anne Vergison, Rianne van Gageldonk-Lafeber, Anne Teirlinck, Jackie Melillo, Tanya Melillo, Piers Mook, Richard Pebody, Ana Paula Coutinho Rehse, Dominique L. Monnet

## Abstract

We assessed the impact of COVID-19 on healthcare workers (HCWs) from data on 2.9 million cases reported from nine countries in the EU/EEA. Compared to non-HCWs, HCWs had a higher adjusted risk of hospitalization (IRR 3.0 [95% CI 2.2-4.0]), but not death (IRR 0.9, 95% CI 0.4-2.0).

**Article Summary Line:** Healthcare workers are hospitalized more frequently than non-healthcare workers when adjusting for age, sex, and comorbidities.

## Introduction

The impact of the ongoing COVID-19 pandemic on healthcare workers (HCWs) has been unprecedented. HCWs are exposed to infected patients making them among the most affected professional groups [1–7], with nurses being the most infected [4]. In a meta-analysis of 97 studies that assessed infection among HCWs, 5% HCW COVID-19 cases had severe complications, and 0.5% died [8]. Using surveillance data provided by European Union and European Economic Area (EU/EEA) countries, we described the risk of disease, hospitalization, intensive care unit (ICU) admission, and death and identified possible risk factors for death in hospitalized HCW and non-HCW (general population) cases of COVID-19.

## Methods

COVID-19 surveillance in the World Health Organization (WHO) European Region is jointly coordinated by the European Centre for Disease Prevention and Control (ECDC) and the WHO Regional Office for Europe. Using case-based surveillance data reported to The European Surveillance System (TESSy), we analyzed the risk of disease, hospitalization, ICU admission, and death among HCWs and non-HCWs using case-to-case study design [8]. Cases with unknown HCW status were assumed to be non-HCWs. All cases with unknown outcome status were recorded as ‘alive’ to avoid overestimating death.

We included all COVID-19 cases aged 20-69 years (working ages) reported to TESSy between January 31, 2020 and January 13, 2021 from countries that had (a) consistent case-based reporting throughout the COVID-19 pandemic (n=15) [9], (b) at least 50% of cases reported as case-based data in TESSy, and (c) at least 40% internal completeness of HCW status [10]. We estimated the crude and adjusted rates for disease, hospitalization, admission to ICU, and death by HCW status using negative binomial regression and accounting for country reporting heterogeneity. We used the total number of HCWs in 2020 by country using the latest HCW population statistics from Eurostat [11] for denominator values in Table 1. We subtracted the HCW population^1^ from the 2020 population statistics from Eurostat [12] to calculate the non-HCW population.

**Table 1.** Number of COVID-19 cases reported from case-based surveillance and COVID-19 attack rate by period, healthcare worker status and outcome, EU/EEA countries†, January 2020 — January 2021*

| Reporting period |  | Reported cases |  |  | Hospitalized cases‡ |  | Cases admitted to ICU‡ |  | Deceased |  |
| --- | --- | --- | --- | --- | --- | --- | --- | --- | --- | --- |
|  |  | Total | HCWs | Non-HCWs§ | HCWs | Non-HCWs§ | HCWs | Non-HCWs§ | HCWs | Non-HCWs§ |
| Overall<br>(Jan 31, 2020 – Jan 13, 2021) | No. | 2,918,100 | 261,080 | 2,657,020 | 8,169 | 119,543 | 716 | 16,946 | 220 | 13,857 |
|  | Attack rate (%) | 4.1 | 10.1 | 3.9 | 0.40 | 0.21 | 0.03 | 0.03 | 0.01 | 0.02 |
| Jan 31, 2020 – May 31, 2020 | No. | 209,993 | 58,734 | 151,259 | 4,557 | 40,139 | 387 | 6,371 | 102 | 6,120 |
|  | Attack rate (%) | 0.3 | 2.3 | 0.2 | 0.22 | 0.07 | 0.019 | 0.011 | 0.004 | 0.009 |
| Jun 1, 2020 – Sep 30, 2020 | No. | 174,072 | 13,435 | 160,637 | 265 | 7,714 | 18 | 673 | 7 | 340 |
|  | Attack rate (%) | 0.2 | 0.5 | 0.2 | 0.01 | 0.01 | 0.001 | 0.001 | 0.000 | 0.001 |
| Oct 1, 2020 – Jan 13, 2021 | No. | 2,534,035 | 188,911 | 2,345,124 | 3,347 | 71,690 | 311 | 9,902 | 111 | 7,397 |
|  | Attack rate (%) | 3.6 | 7.3 | 3.5 | 0.16 | 0.13 | 0.015 | 0.017 | 0.004 | 0.011 |

\*EU/EEA, European Union and Economic Area; HCWs, healthcare workers; ICU, intensive care unit; TESSy, The European Surveillance System.
†Nine EU/EEA countries with consistent reporting case-based surveillance data reported to TESSy for cases aged 20-69 years: Austria, Finland, Ireland, Italy, Luxembourg, Malta, the Netherlands, Norway, and Slovakia. Consistent reporting was defined as reporting case-based surveillance data throughout the entire study period of January 2020-January 2021.
‡One country (the Netherlands) was excluded as data on hospitalizations and ICU admissions were incomplete or not reported. §Non-HCWs include cases with unknown HCW status.

As HCWs are tested more frequently than non-HCWs (risk of detection bias), we estimated incidence rate ratios (IRRs) and 95% confidence intervals (CIs) for death for only hospitalized cases using negative binomial regression, adjusting for age group (20-29, 30-39, 40- 49, 50-59, and 60-69 years), sex, comorbidities^2^, and three reporting periods (January 31 to May 31, 2020, June 1 to September 30, 2020, and October 1, 2020 to January 13, 2021). We included the time variable in the model to control for differences in testing, reporting, and healthcare capacity during the pandemic. To account for country reporting heterogeneity, we calculated adjusted IRRs using robust clustered standard errors with reporting countries as a cluster effect in the negative binomial regression model. The level of statistical significance was p<0.05 and the analyses were performed using Stata 16 and R 3.6.2.

## Results

Nine countries met our inclusion criteria: Austria, Finland, Ireland, Italy, Luxembourg, Malta, the Netherlands, Norway, and Slovakia. These countries (70 million total population aged 20-69, 2.6 million HCWs) reported a total of 2.9 million cases from January 31, 2020 to January 13, 2021, corresponding to an overall attack rate of 4.1%. The proportion of HCW cases in the first reporting period (28%) differed greatly from reporting periods later in the pandemic (7.7%-7.5%) (Table 1). The attack rate in the population aged 20-69 was 10.1% for HCWs and 3.9% for non-/unknown-HCWs (Table 1), corresponding to a crude IRR of 2.6 and an adjusted IRR of 3.0 (95% CI 2.2-4.0) for HCWs. The adjusted risk of COVID-19 requiring hospitalization or admission to ICU was respectively 1.8 and 1.9 times higher in HCWs than in non-HCWs (95% CIs 1.2-2.7 and 1.1-3.2, respectively), but the adjusted risk of death was not significantly different (adjusted IRR 0.9, 95% CI 0.4-2.0).

Of 127,712 hospitalized cases aged 20-69 years, 8,169 (6.4%) were HCWs and 119,543 (93.6%) non-HCWs (Table 1). Of those hospitalized, 147 (1.8%) HCWs died, compared to 9,773 (8.2%) non-HCWs (Table 2).

**Table 2.** Number of hospitalized COVID-19 cases and risk of death (IRR) by characteristics of cases, EU/EEA countries†, January 31, 2020-January 13, 2021*

| Characteristic | No. hospitalized cases<br>(% within stratum) |  | Number of deaths (%) |  | IRR | 95% CI |
| --- | --- | --- | --- | --- | --- | --- |
|  | HCWs | Non-HCWs | HCWs | Non-HCWs |  |  |
| HCW and ICU admission |  |  |  |  |  |  |
| Non-HCW, not admitted to ICU | -- | 102,597 (80.3) | -- | 4,823 (4.7) | Ref. | -- |
| Non-HCW, admitted to ICU | -- | 16,946 (13.3) | -- | 4,950 (29.2) | 4.6 | 2.7-7.7 |
| HCW, not admitted to ICU | 7,453 (5.8) | -- | 48 (0.6) | -- | 0.30 | 0.2-0.6 |
| HCW, admitted to ICU | 716 (0.6) | -- | 99<br>(13.8) | -- | 0.62 | 0.4-0.9 |
| Sex |  |  |  |  |  |  |
| Female | 4,786 (58.6) | 43,729 (36.6) | 34 (0.7) | 2 537 (5.8) | Ref. | -- |
| Male | 3,382 (41.4) | 75,808 (63.4) | 113<br>(3.3) | 7 236 (9.5) | 1.3 | 1.1-1.5 |
| Age group, y |  |  |  |  |  |  |
| 20-29 | 518 (6.3) | 6,673 (5.6) | 0(0.0) | 44 (0.7) | Ref. | -- |
| 30-39 | 1,099 (13.5) | 10,724 (9.0) | 2 (0.2) | 149 (1.4) | 1.6 | 1.0-2.6 |
| 40-49 | 1,966 (24.1) | 18,918 (15.8) | 12 (0.6) | 561 (3.0) | 2.1 | 1.3-3.2 |
| 50-59 | 3,027 (37.1) | 36,398 (30.4) | 42 (1.4) | 2,274 (6.2) | 3.0 | 2.0-4.6 |
| 60-69 | 1,559 (19.1) | 46,830 (39.2) | 91 (5.8) | 6,745 (14.4) | 5.8 | 3.8-8.8 |
| Any comorbidity |  |  |  |  |  |  |
| No | 6,654 (81.5) | 95,485 (79.9) | 89 (1.3) | 6,347 (6.6) | Ref. | -- |
| Yes‡ | 1,515 (18.5) | 24,058 (20.1) | 58 (3.8) | 3,426 (14.2) | 2.0 | 1.8-2.3 |
| Reporting period |  |  |  |  |  |  |
| Jan 31, 2020 – May 31, 2020 | 4,557 (55.8) | 40,139 (33.6) | 83 (1.8) | 4,520 (11.3) | Ref. | -- |
| Jun 1, 2020 – Sep 30, 2020 | 265 (3.2) | 7,714 (6.5) | 4 (1.5) | 212 (2.7) | 0.53 | 0.42-0.67 |
| Oct 1, 2020 – Jan 13, 2021 | 3,347 (41.0) | 71,690 (60.0) | 60 (1.8) | 5,041 (7.0) | 0.83 | 0.71-0.96 |

\*EU/EEA, European Union and European Economic Area; HCWs, healthcare workers;
IRR, incidence rate ratio; CI, confidence interval.
†Eight EU/EEA countries with consistent reporting case-based surveillance data reported to TESSy for hospitalized cases aged 20-69 years, including ICU admission: Austria, Finland, Ireland, Italy, Luxembourg, Malta, Norway, and Slovakia. Consistent reporting was defined as reporting case-based surveillance data throughout the entire study period of January 2020-January 2021.
‡Presence of at least one of the following: asthma, cancer, cardiac disease, diabetes, HIV,
hypertension, kidney disease, liver disease, lung disease, neuromuscular disease, obesity, pregnancy, smoking status, tuberculosis, and other/unspecified.

Hospitalized HCWs had a lower risk of in-hospital death (IRR 0.3 [95% CI 0.2-0.6]), but this effect was halved for HCWs admitted to an ICU (IRR 0.6 [95% CI 0.4-0.9]) (Table 2).

ICU admission was an independent predictor of death after hospitalization (IRR 4.6 [95% CI 2.7-7.7]), which likely reflects risk factors for severity of disease that were not included in the model (Table 2). Being 60-69 years old compared to cases aged 20-29 (IRR 5.8 [95% CI 3.8- 8.8]), having at least one comorbidity (IRR 2.0 [95% CI 1.8-2.3]) or being male (IRR 1.3 [95% CI 1.1-1.5]) was associated with increased risk of death among hospitalized cases (Table 2). Being hospitalized between June-September 2020 was associated with a decreased risk of death (IRR 0.5 [95% CI 0.4 to 0.7]) compared to the early phase of the pandemic. There was no increased risk of death based on the number of comorbidities for either hospitalized HCWs or hospitalized non-HCWs.

## Discussion

In our analyses, HCWs in all countries were at increased risk of COVID-19- related hospitalization, suggesting an increased risk of exposure to SARS-CoV-2. However, HCWs may seek and receive care earlier than non-HCWs, thereby impacting hospitalization and survival rates. By restricting analyses to cases requiring hospitalization, we limited the effect of differential underdiagnosis of cases in non-HCWs vs. HCWs who are regularly tested in healthcare settings.

HCWs were less likely than non-HCWs to die following hospitalization. Risk factors for death such as sex, age, and comorbidity reflect what has been previously reported [13]. After May 31, 2020, there was a lower risk of dying in the hospital for all COVID-19 cases than during the initial phase of the pandemic. Testing practices changed over time and the availability of better equipment, knowledge, and treatment of cases most likely influenced health outcomes for all cases. During the second and third reporting periods, the decreased risk of death may be linked to improved management of severe cases [14].

In addition, it is plausible that HCWs, because they are close to the healthcare system, receive early treatment and are better able to identify clinical symptoms that can lead to severe outcomes that affected their survival rates. HCWs in our analyses may also have benefited from the healthy worker effect [15] or HCWs with high-risk comorbidities may have left the practice during the pandemic.

While we tried to control for detection bias by restricting our multivariable analysis to hospitalized cases, it is possible that HCWs were hospitalized with less severe illness than non-HCWs. Country-specific definitions of HCWs vary and we did not have data on the professions of HCW cases, so their distribution and data completeness in our surveillance dataset is unknown. While misclassification of cases (either as non-HCWs or as alive) was possible, the adjusted IRRs did not change when we restricted the analysis to only cases with known HCW and outcome statuses.

In conclusion, HCWs were at higher risk for COVID-19-related hospitalization than non-HCWs, which could possibly be explained by their proximity to healthcare services and prompt recognition of illness. Among hospitalized cases, the risk of death was lower for HCWs than for non-HCWs, likely due to the healthy worker effect, better or earlier access to treatment, and under ascertainment. Further research is needed on exposure levels by HCW profession to fully explore the risk factors for COVID-19-related hospitalization and death among HCWs.

## Data Availability

Data are publicly available through https://www.ecdc.europa.eu

## Acknowledgments

We would like to gratefully acknowledge all the contributing public health practitioners who provided data used in this manuscript. We would also like to acknowledge the contributions of Gianfranco Spiteri, Nick Bundle, Pasi Penttinen, Andrew J Amato-Gauci, Karl Ekdahl, Antonino Bella, and all ECDC employees and staff who helped collect, analyze, and report these data during the public health emergency response. We would like to acknowledge the contributions of all public health practitioners in EU/EEA countries conducting surveillance and reporting data related to COVID-19. ECDC continues to collect surveillance data and collaborate with the WHO to monitor risk factors and advise EU/EEA countries on the severity of disease as the pandemic progresses.

## Disclaimers

The authors alone are responsible for the views expressed in this publication and they do not necessarily represent the decisions or policies of the institutions with which they are affiliated. Data are publicly available through https://www.ecdc.europa.eu.

## Footnotes

^1^ HCW professional categories included: (i) medical doctors, (ii) nursing and midwifery, (iii) dentistry, (iv) pharmaceutical, (v) environmental and occupational health and hygiene personnel, medical and pathology laboratory personnel, (vii) physiotherapy personnel, (viii) traditional and complementary medicine personnel, and (ix) community HCWs.

^2^ Comorbidities included asthma, cancer, cardiac disease, diabetes, HIV, hypertension, kidney disease, liver disease, lung disease, neuromuscular disease, obesity, pregnancy, smoking status, tuberculosis, and other/unspecified.

